# Sex Moderates the Relationship that Number of Professional Fights has with Cognition and Brain Volumes

**DOI:** 10.1101/2020.06.19.20135897

**Authors:** Lauren L. Bennett, Steve Stephen, Charles Bernick, Guogen Shan, Sarah J. Banks

## Abstract

**Objective:** Incidence of concussions and report of symptoms are greater amongst women across sports. While structural brain changes and cognitive declines are associated with repetitive head impact (RHI), the role of sex is not well understood. This study aimed to determine if there is a moderating effect of sex on the relationship number of professional fights has with cognitive functioning and regional brain volumes in a cohort of boxers, mixed martial artists, and martial artists.

**Methods:** 55 women were matched with 55 men based on age, years of education, ethnicity, and fighting style. Cognition was assessed via CNS Vital Signs computerized cognitive battery and supplemental measures. Structural brain scans, demographic data, and number of professional fights (NoPF) were also considered. Matched pairs were compared via analysis of covariance, accounting for total brain volume. Within-subject moderation models were utilized to assess the moderating effect of sex on the relationship between NoPF and brain volumes and cognitive performance.

**Results:** Men were observed to have poorer performance on measures of psychomotor speed when compared to women. On a series of analyses assessing the role of sex as a moderator of the relationship between NoPF and regional brain volumes/cognitive performance, a significant moderation effect was observed across multiple measures of cognitive functioning, such that men had poorer performance. Differences in numerous regional brain volumes were also observed, such that the relationship between NoPF and brain volumes was steeper amongst men.

**Conclusion:** Sex was observed to be an important moderator in the relationship between NoPF, aspects of cognitive functioning, and volumes of numerous brain regions, suggesting that sex differences in neuroanatomic and cognitive response to RHI deserve further attention.

## INTRODUCTION

Multiple epidemiological studies have demonstrated that male athletes have been shown to be at lower risk for sports-related concussion than female athletes within both practice and competition across numerous sports (Merritt, Padgett, & Jak, 2019). Specifically, women participating in basketball, soccer, lacrosse, softball/baseball, tennis, outdoor track, and cross-country are at increased risk for sustaining a concussion than men (Lincoln et al., 2011; Marar, McIlvain, Fields, & Comstock, 2012; Zuckerman et al., 2015). Moreover, following head injury, female athletes have been found to endorse greater severity of concussion-related symptoms, demonstrate greater declines in cognitive functioning, and slower physiological recovery post-injury. Prior research suggests there are sex differences in the changes to both structural and metabolic brain functioning following concussions for athletes participating in other sports (i.e., ice hockey, soccer, water polo; Chamard, Lefebvre, Lassonde, & Theoret, 2016; Covassin, Elbin, Bleecker, Lipchik, & Kontos, 2013; Covassin, Moran, & Elbin, 2016; Fakhran, Yaeger, Collins, & Alhilali, 2014; Frommer et al., 2011; Sollmann et al., 2018). Very little research has been completed in sex differences in male and female professional fighters (Merritt et al., 2019)

In addition, recent professional fight records comparing fight outcomes across men and women may be an indication of different rates of repetitive head impact exposure, suggesting need for further exploration of sex differences. When examining Nevada state fight records from 2015 through 2017, it appears women are less likely to be knocked out (KO) or sustain a technical knockout (TKO) than male fighters, overall (please see Table 1; Nevada State Athletic Commission, n.d.). This data is in contrast to the higher rate of concussions observed amongst female athletes who participate in most other sports. Notably, as fewer women compete in combat sports, it is difficult to determine if the same trend would exist if the samples were to be equal sized. To date, the relationship between sex, regional brain volumes, and cognitive performance amongst male and female professional fighters has not been examined. This study aimed to investigate if there is a differential effect of sex on the relationship that number of professional fights (NoPF) has with cognitive functioning and brain region volumes in professional fighters.

**Table 1.** 2015 to 2017 Nevada commission fight records

|  | KO | TKO | Decision/Draw | Submission | Total (%<br>KO/TKO) |
| --- | --- | --- | --- | --- | --- |
| Boxer | 34 | 170 | 296 | N/A | 500 (41) |
| Men | 33 | 168 | 288 | - | 489 (41) |
| Women | 1 | 2 | 8 | - | 11 (27) |
| MMA | 14 | 136 | 263 | 137 | 550 (27) |
| Men | 12 | 123 | 225 | 107 | 467(29) |
| Women | 2 | 13 | 38 | 30 | 83 (18) |
| Martial Arts | 4 | 14 | 34 | N/A | 52 (35) |
| Men | 4 | 12 | 32 | - | 48 (33) |
| Women | 0 | 2 | 2 | - | 4 (50) |

Note: Decision/Draw includes any result labeled as unanimous decision, split decision, draw, or majority decision. Submission includes any result labeled as choke outs, tap out, or submission. Outcomes of fights that were later changed to “no decision” or labeled as “cancelled” were not included in this count.

## METHODS

### Study design

Participants were drawn from an ongoing longitudinal observational study of professional combat sport athletes, the Professional Fighters Brain Health Study (PFBHS; Bernick et al., 2013). As part of the PFBHS, participants are assessed at baseline and at subsequent one-year intervals. For the purposes of this study, baseline imaging, demographic, number of professional fights, and cognitive functioning data were considered. Given the small number of women with longitudinal data, we restricted the current analysis to the baseline data, thus we report cross-sectional analyses here. While collected, self-report of number of amateur fights and concussions were excluded from analyses as these data reflect high levels of variability and are uncorroborated. Since the number of professional fights (NoPF) could be validated, it was used as a proxy for repetitive head impact exposure with consideration of training exposure, in addition to exposure during the actual match.

The study was approved by the local institutional review board (#10-944) and written informed consent was obtained from all study participants. Study visits for all participants included in these analyses were performed at the Cleveland Clinic Lou Ruvo Center for Brain Health in Las Vegas, Nevada between 03/30/2011 and 10/18/2017. For a detailed explanation of study methods see Bernick et al (2013).

### Participants

The study cohort consisted of all 55 women fighters enrolled in the PFBHS to date and 55 male fighters who were matched with the female fighters on the following variables: age, years of education, ethnicity, and type of competitive fighting, including boxing, mixed martial arts, and martial arts (e.g. kickboxing, Muay Thai, judo). Both retired and active professional fighters were included in the analyses for this study. Participants were not seen for their baseline or follow up visits for at least 45 days following their most recent fight.

### Cognitive and psychological assessment

Participants completed a short battery of computerized cognitive and motor tests including symbol digit coding, finger-tapping, and Stroop-like tasks from the CNS Vital Signs program (Gualtieri & Johnson, 2006). Performance across CNS Vital Sign subtests produced composite score of the following domains: Verbal Memory (word list recognition), Processing Speed (the number of correctly completed items on symbol digit coding, while accounting for incorrect responses), Psychomotor Speed (bilateral finger-tapping speed and number correct on a digit symbol digit coding task), and Reaction Time (response time on Stroop-like tasks). Prior research has found that men perform significantly better on right-sided finger-tapping, but no other significant sex-based performance differences were observed (Iverson, Brooks, & Ashton Rennison, 2014).

Beyond the CNS Vital Signs battery, fighters completed six additional measures of cognitive functioning during assessment, including supplemental measures of processing speed (a timed reading passage and a computerized version of Trails A via the iCOMET battery [speeded connection of numeric dots]), language (semantic fluency [ability to name items belonging to given a semantic category], a word task [ability to pronounce words with irregular, non-phonetic spellings]), executive functioning (letter fluency [ability to generate words beginning with given letter], and a computerized version of Trails B via the iCOMET battery [speeded, alternating connection of numeric and alphabetic dots]).

### Imaging

Brain MRI scans were conducted on a MAGNETOM Verio 3-tesla scanner (Siemens Medical Systems, AG, Erlangen, Germany) with volumetric values derived from T1-weighted images via FreeSurfer, version 6 (Fischl, 2012).

Volumetric segmentation was completed on the MPRAGE sequence using the Freesurfer Version 6.0 image analysis suite. Conventional sagittal 3D magnetization-prepared rapid acquisition with gradient echo (MPRAGE) T1 (voxel size = 1 × 1 × 1.2 mm; flip angle/repetition time [TR]/echo time [TE]/inversion time [TI] = 9/2300/2.98/900 ms; scan time = 9:14), axial turbo spin-echo (TSE) T2 (voxel size = 0.8 × 0.8 × 4 mm; TR/TE =5000/84 ms; 38 slices; scan time = 0:57), axial TSE fluid-attenuated inversion recovery (voxel size = 0.8 × 0.8 × 4 mm; TR/TE/TI = 7000/81/2220 ms; 38 slices; scan time = 2:36), and axial susceptibility weighted imaging (SWI) (voxel size = 0.9 × 0.9 × 0.9 mm; TR/TE = 20/27 ms; 36 slices; scan time = 1:17) sequences were used for this study.

Volumetric segmentation was performed on the MPRAGE sequence using the Freesurfer Version 6.0 image analysis suite (http://surfer.nmr.mgh.harvard.edu/). Procedures for measuring cortical thickness have been validated against histological analysis and manual measurements. Freesurfer morphometric procedures have demonstrated good test-retest reliability across scanner manufacturers and across field strengths.

### Analyses

Guided by prior findings from the PFBHS, as well as other studies highlighting structural and metabolic changes following repetitive head impact, the following brain regions were chosen *a priori* for examination: the putamina, hippocampi, amygdalae, caudates, and thalami (Bernick & Banks, 2013; Bernick et al., 2015; Chamard et al., 2016; Lee, Bennett, Bernick, Shan & Banks, 2019; Schultz et al., 2018). When applicable, both left and right sides of brain regions were evaluated. Additionally, total brain volume was controlled for in all imaging analyses. Moderation analyses were conducted in SAS, version 9. All other analyses were conducted in SPSS version 23.

Analysis of variance tests were run to examine group differences between female and male fighters on the four CNS Vital Signs composite scores (Verbal Memory, Processing Speed, Psychomotor Speed, Reaction Time), as well as supplemental measures of processing speed (timed reading task, computerized version of Trails A), language (semantic fluency), and executive functioning (letter fluency, computerized version of Trails). Analysis of covariance tests were run to examine group differences across regional brain volumes between male and female fighters, while accounting for total brain volume. To assess the moderating role of sex in the relationship between fight exposure and regional brain volumes and cognitive performance, a series of within-subject moderation models were computed.

Twenty models were computed, one for each of the five brain regions identified *a priori* and each of the 10 cognitive outcome measures. Sex as a moderator within models was considered at *p <* .05. Exploratory analyses to characterize models were considered at *p <* .05.

## RESULTS

### Demographic data

Fighters ranged in age from 19 to 55 years, with a mean age of 30.70 (SD = 6.7). Of the 110 fighters, 101 were active fighters consisting of 52 men and 49 women. Of the 9 retired fighters, 3 were men and 6 were women. Participants were encouraged to select all race categories that applied to them. Self-defined race was 69% (n = 76) white; 13% (n = 14) black; 8% (n = 9) other; 6% (n = 6) Pacific Islander; 3% (n = 3) Asian. Four participants (n =4) did not provide their race. The majority of fighters identified as mixed martial artists (MMAs; n = 64). The mean number of years of education completed was 14.46 (SD = 2.0). As participants were matched, there were no significant differences between male and female fighters with regard to age, years of education, ethnicity, and type of fighting (e.g., boxer, MMA, MAs; all *p*’s > .2). Fighters had a mean of 12.8 professional fights (SD = 15.8). Female and male fighters did not significantly differ in the number of professional fights fought (*p* = .764). Additional demographic data are presented in Table 2.

**Table 2.** Participant demographic data.

|  | Fighters (n = 110) | Male fighters (n = 55) | Female fighters (n = 55) |
| --- | --- | --- | --- |
| Age (mean [SD]) | 30.7 (6.7) | 30.9 (6.9) | 30.6 (6.6) |
| Education (mean [SD]) | 14.0 (3.7) | 13.9 (3.7) | 14.2 (3.8) |
| Number of professional fights (mean [SD]) | 12.8 (15.8) | 12.4 (12.6) | 13.3 (18.5) |
| Active fighters | 101 | 49 | 52 |
| Type of fighting |  |  |  |
| Boxers (%) | 32 (29.1) | 15 (27.3) | 17 (30.9) |
| MMA's (%) | 69 (62.7) | 38 (69.1) | 31 (56.4) |
| MAs (%) | 9 (8.2) | 2 (3.6) | 7 (12.7) |
| Years of education completed | 108 |  |  |
| Middle school (%) | 1 (0.9) |  |  |
| Some high school (%) | 2 (1.9) |  |  |
| High school (%) | 22 (20.4) |  |  |
| Some college (%) | 8 (7.4) |  |  |
| Associate's degree (%) | 31 (28.7) |  |  |
| Bachelor's degree (%) | 36 (33.3) |  |  |
| Some graduate school (%) | 1 (0.9) |  |  |
| Master's degree (%) | 6 (5.6) |  |  |
| Doctoral degree (%) | 1 (0.9) |  |  |
| Number of professional fights |  |  |  |
| 0 (%) |  | 21 (19.1) |  |
| 1-10 (%) |  | 39 (35.5) |  |
| 11-20 (%) |  | 27 (24.6) |  |
| 21-30(%) |  | 13 (11.8) |  |
| 31-40 (%) |  | 5 (4.5) |  |
| 41-50 (%) |  | 0 |  |
| 51-60 (%) |  | 3 (2.7) |  |
| 61 or more (%) |  | 2 (1.8) |  |

Note: As male and female fighters were matched, there were no significant differences between male and female fighters with regard to age, years of education, ethnicity, and type of fighting (e.g., boxer, MMA, MAs; all $P$ 's > .2)

### Sex differences in regional brain volumes

ANCOVAs were conducted to assess differences in regional brain volumes while controlling for total brain volume. Female fighters (mean = 1624.18) differed significantly from male fighters (mean = 1669.05) in left amygdala (*F*(1, 106) = 8.417; *p =* .005), when accounting for total brain volume. In contrast, the right amygdala, bilateral hippocampi, thalamus, and caudate volumes were not significantly different as a function of sex (all *p*’s >.05). These results suggest that female fighters have larger left amygdala than their male fighter counterparts.

### Sex differences in cognitive performance

Male fighters had poorer performance on CNS Vital Signs Psychomotor Speed measures (*F*(1, 106) = 8.32; *p* = .005). There were no significant differences on CNS Vital Signs composite scores of Verbal Memory, Processing Speed, and Reaction Time (all *p*’s > .05). Similarly, there were no significant difference on supplement cognitive functioning measures of processing speed (Trails A and passage reading time), language (semantic fluency and word reading), and executive functioning (Trails B and letter fluency) performance as a function of sex (all *p*’s > .05). All analyses accounted for education.

### Sex as a moderator of the relationship between fight exposure and regional brain volumes

In order to explore the role of sex as a moderator of the relationship between fight exposure and regional brain volumes, a series of within-subject moderation analyses were computed according to the above-stated parameters. A significant moderation effect of sex was observed on right hippocampus, right thalamus, left putamen, left amygdala, and right amygdala (please see Figure 1A-E and Table 3). While smaller right hippocampus, right thalamus, left putamen, and bilateral amygdala was associated with greater fight exposure in both men and women, the relationship between number of professional fights and volumes was much steeper amongst men than the relationship between number of professional fights and volumes amongst women.

**Table 3.** Moderation effect of sex on subcortical regional brain volumes

|  | *P*<br>value | *b* | 95% confidence<br>interval | Slope of male<br>fighters | Slope of<br>female fighters |
| --- | --- | --- | --- | --- | --- |
| Right<br>hippocampus | 0.015 | -6.10 | -10.95 to -1.25 | -15.37 | - 3.17 |
| Right thalamus | 0.020 | -13.45 | -24.70 to -2.21 | -30.60 | - 3.69 |
| Left putamen | 0.046 | -8.74 | -17.30 to -0.18 | -17.48 | -2.99 |
| Left amygdala | 0.002 | -4.79 | -7.77 to -1.82 | -10.53 | -0.94 |
| Right amygdala | 0.007 | -4.21 | -7.20 to -1.21 | -9.06 | -0.65 |
| Right putamen | 0.015 | -8.68 | -10.95 to -1.25 | -16.62 | 0.73 |

**Figure 1.**
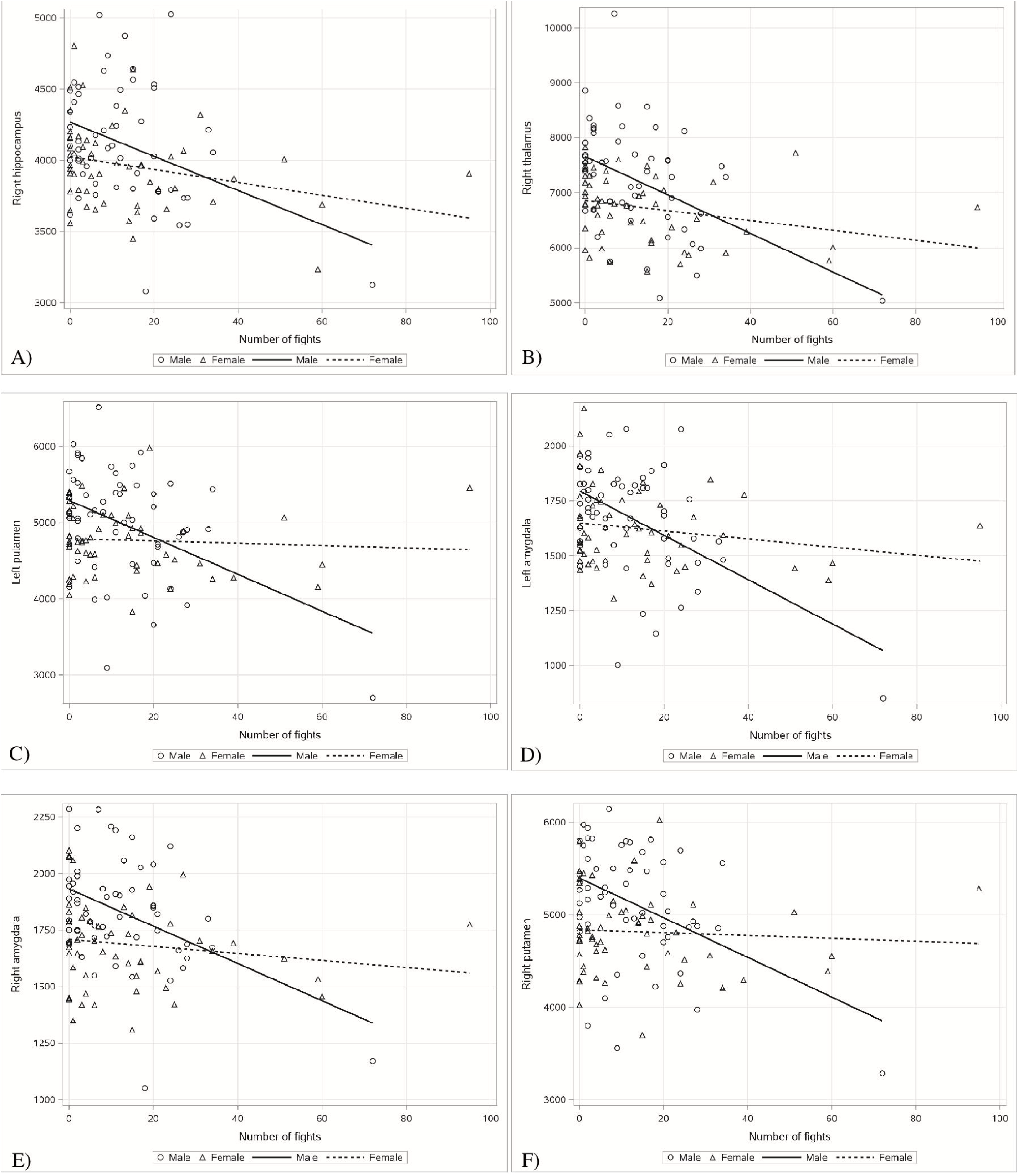
Relationship between number of professional fights and subcortical regional brain volumes (mm^3) by sex: A) right hippocampus, B) right thalamus, C) left putamen, D) left amygdala, E) right amygdala, and F) right putamen

While a significant moderation effect of sex was also observed on right putamen, the relationship amongst women and men differed. Amongst male fighters, higher number of professional fights was associated with smaller right putamen volume. In contrast, greater right putamen volume was associated with higher number of professional fights amongst female fighters (please see Figure 1F and Table 3). The left hippocampus, left thalamus, and bilateral caudate were not found to be significant moderators in the relationship between number of professional fights and regional brain volumes (*p >* .05).

### Sex as a moderator of the relationship between fight exposure and cognitive performance

A significant moderation effect of sex was also observed on CNS Vital Signs Verbal Memory and Reaction Time performance. While lower Verbal Memory performance was associated with a higher number of professional fights in male fighters, Verbal Memory performance was positively associated with number of professional fights amongst female fighters (please see Figure 2A and Table 4). Looking at CNS Vital Signs Reaction Time performance, which is reverse scored with lower scores equating to a faster, better performance, while the Reaction Time performance of both male and female fighters was found to be better with greater number of professional fights, the relationship between number of professional fights and Reaction Time performance was much steeper amongst men than the relationship between number of professional fights and Reaction Time performance amongst women (please see Figure 2B and Table 4).

**Table 4.** Moderation effect of sex on subcortical regional brain volumes

|  | *p* value | *b* | 95% confidence interval | Slope of male fighters | Slope of female fighters |
| --- | --- | --- | --- | --- | --- |
| CNS Vital Signs Verbal Memory | <0.0001 | -0.23 | -0.33 to -0.14 | -0.34 | 0.1262 |
| CNS Vital Signs Reaction Time | 0.046 | 2.23 | -0.037 to 4.42 | 5.33 | 0.8708 |

**Figure 2.**
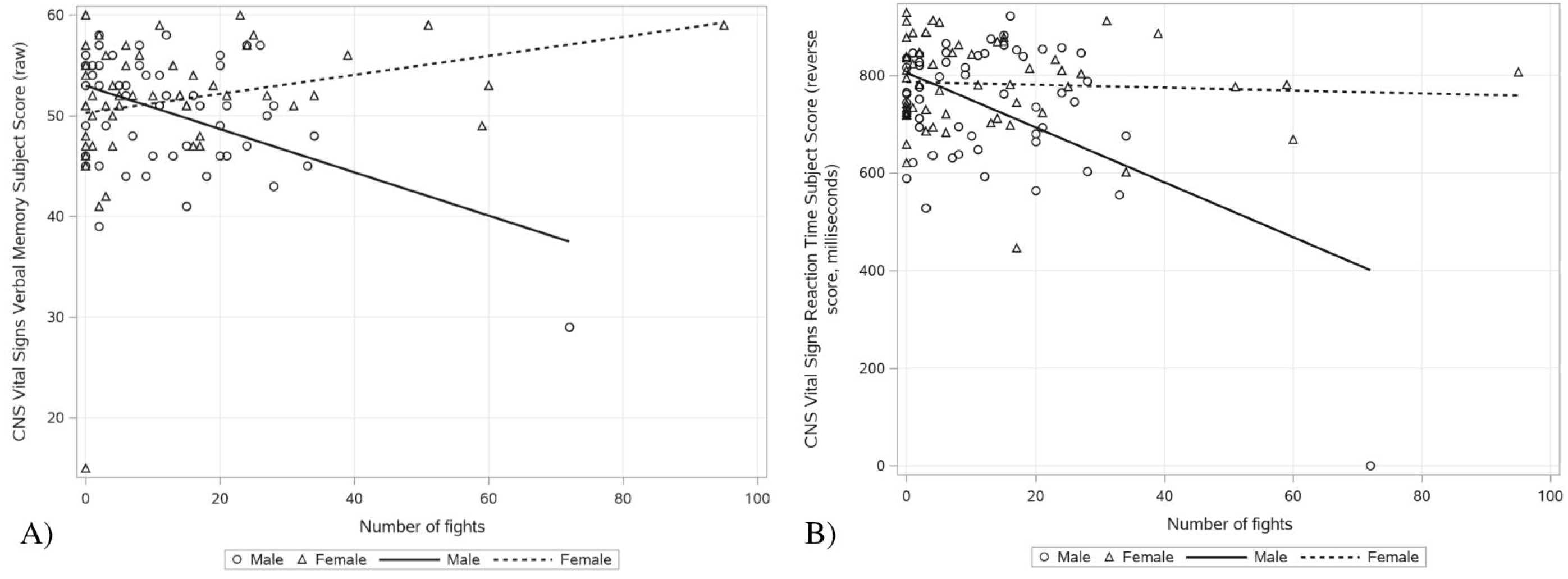
Relationship between number of professional fights and CNS Vital Signs performance by sex: A) Verbal Memory Subject Score (raw number correct) B) Reaction Time Subject Score (reverse scored; milliseconds) *Note:* As CNS VS Reaction Time is reverse scored, graph depicts maximum score – raw score for each participant and is reported in milliseconds

Sex was not found to be a significant moderator in the relationship between number of professional fights and performance on measures of processing speed (CNS Vital Signs Processing Speed, Trails A, timed reading task), psychomotor speed (CNS Vital Signs Psychomotor Speed), language (semantic fluency, word reading task), or executive functioning (letter fluency, Trails B; all *p’s* >.05).

## DISCUSSION

The current findings reveal key insights into sex-based differences. When accounting for whole brain volume, women were found to have larger left amygdala than men. While prior research is equivocal, a recent meta-analysis suggests the amygdalae are not sexually dimorphic (Marwha, Halari, & Eliot, 2017). Additional sex-based differences emerged when exploring the role of sex as a moderator of the relationship between fight exposure and regional brain volumes or cognitive performance.

With regard to regional brain volumes, relevant subcortical smaller volumes were associated with greater number of professional fights amongst both male and female fighters. Notably, the relationship between number of professional fights and regional brain volumes were observed to be much steeper in men. Interestingly, while a significant moderation effect of sex was observed on right putamen such that lower volumes in men were associated with higher number of professional fights, an inverse relationship was observed amongst women. Notably, a level of noise exists in the measurement of regional brain volumes via MRI. Prior research has found that the test-retest differences of structures is up to five percent, so the association between number of professional fights and larger volumes in the right putamen may reflect the noise of the measure itself (Iscan et al., 2015). Alternatively, as the putamen is a highly connected brain region, it may be that as volumes of other surrounding regions are decreasing, the volume of the putamen increases in an effort to compensate. In addition, prior research suggests that the basal ganglia may be one of the earliest brain regions to manifest inflammation in some disease processes (e.g., HIV; Wright et al., 2016). It may be that the slight observed “growth” in the right putamen is actually a reflection of inflammation in response to exposure to repetitive head impact.

Sex also moderated the relationship between number of professional fights and two aspects of cognitive functioning, Verbal Memory and Reaction Time. Notably, a greater number of professional fights was associated with poorer Verbal Memory performance amongst male fighters, while an inverse relationship was observed amongst female fighters. Recent research has demonstrated that factors impacting hormones (e.g., use of hormonal contraceptives) in women younger than the age of 65 can have a profound impact on cognitive functioning, with the strongest association between verbal memory and hormonal contraceptive use (Maki & Sundermann, 2009). As age was observed to be highly collinear with number of professional fights, it may be that as number of professional fights and age increased, so too does opportunity for increased hormonal contraceptive use amongst female fighters. Another study has shown how pre-menopausal women (mean age = 24.2) taking oral contraceptives had improved verbal memory compared to women not taking oral contraceptives (mean age = 25.69), which may also contribute to the inverse relationship discovered in our female cohort (mean age = 31; Mordecai, Rubin, & Maki, 2008). In contrast, the poorer Reaction Time performance amongst both male and female fighters was associated with higher number of professional fights. Notably, the relationship between Reaction Time performance and number of professional fights was much steeper amongst men.

While smaller regional brain volumes and poorer cognitive performance was largely associated with higher number of professional fights in both men and women, female fighters were consistently less negatively impacted than male fighters. This may reflect lower velocity punches, greater resilience, or a combination of multiple factors. For example, Kimm and Thiel (2015) found that while velocity of punches increases with experience (i.e. more fights) for all fighters, regardless of gender, women have lower velocity punches than men at all levels of experience. These findings also suggest as NoPF rise, so too does velocity of punches and, subsequently, risk of head injury to the fighter’s opponent. Another recent study examined backward arm cranking power output as a proxy for punch power output, demonstrating that men’s “muscle performance for protracting the arm to propel the first forward” was significantly greater than their female counterparts’ performance (Morris, Link, Martin, & Carrier, 2020). The described discrepancy in punch velocity may also explain why female fighters appear to show less delirious cognitive and regional brain volume impact.

The current findings are further supported by prior PFBHS research outcomes that demonstrate volumetric reduction and negative impacts on cognitive performance with increased head injury exposure (i.e., increased NoPF). One study using NoPF and years of fighting as a proxy for head injury exposure, found increased exposure was associated with decreased thalamic and caudate volumes, as well as poorer processing speed performance (Bernick 2015). Similarly, when comparing boxers, mixed martial artists, and martial artists, fighting style was observed to moderate the relationship between NoPF, cognitive performance, and regional brain volumes (Stephen, 2019).

Though the PFBHS is highly unique in that the data reflects a large cohort of professional fighters, various limitations of the current study must be considered. Less than 10 percent of the entire PFBHS sample is female. As the cohort of female fighters is significantly smaller than the cohort of male fighters, only baseline data could be considered in an effort to retain as many participants as possible and preserve statistical power. Notably, as the cohort of martial artists practice a wide variety of fighting styles (e.g., kickboxing, Muay Thai, taekwondo, and jiu-jitsu), the martial arts cohort may reflect a wide range of exposure to repetitive head impact. Similarly, number of professional fights (NoPF) only accounts for professional matches. As such, the impact of training for and participating in amateur matches is not considered. Additionally, rather than considering KO/TKO from professional matches alone, NoPF was utilized as a proxy for head impact exposure (inclusive of subconcussive and concussive impacts) across the period of training for and participating in professional level matches. Moreover, fighters’ weight classes were not considered, but may impact the incidence and severity of repetitive head impact. As approximately 40 analyses (some including covariates and multiple variables) were completed, an increased potential for false positives is acknowledged. One counterintuitive finding (i.e. larger putamen volumes among women with more exposure) was observed, indicating additional research is required. Finally, additional, unconsidered factors (i.e., drug use, socioeconomic status, genetic predisposition) may contribute to declines in cognitive functioning and regional brain volumes, rather than exposure to repetitive head impact alone.

As collection of data is ongoing, further analyses exploring changes over time when accounting for sex differences may provide insight into longer term discrepancies in regional brain volumes and cognitive functioning. These baseline results do not necessarily imply differences in long-term impacts of repetitive head impact as a function of sex, but longitudinal data over time comparing male and female fighters may have predictive value. Comparing male and female retired fighters may also provide some information about the long-term concussive impact differences.

In summary, this study adds to our understanding that sex-based cognitive and volumetric differences in response to repetitive head impact exist amongst fighters. The main finding is that sex is an important moderator in the relationship between number of professional fights, aspects of cognitive functioning, and brain volumes of numerous regions. While smaller regional brain volumes and poorer cognitive performance were generally associated with greater number of professional fights amongst men and women, female fighters were consistently less negatively impacted than male fighters.

## Data Availability

The data will be made available on a case-by-case basis, upon request, without undue objections.

## ACKNOWLEDGEMENTS

Conflicts of Interest None

